# Study protocol for a cross-sectional study on knowledge, attitude, and practice towards thalassemia among Indonesian youth

**DOI:** 10.1101/2021.04.11.21255264

**Authors:** Muhammad Maulana Wildani, Visabella Rizky Triatmono, Edward Christopher Yo, Mikhael Yosia, Pustika Amalia Wahidiyat

**Author notes:** ***Corresponding author*** Pustika Amalia Wahidiyat, Division of Hematology and Oncology, Department of Child Health, Cipto Mangunkusumo Hospital - Faculty of Medicine Universitas Indonesia, Jalan Pangeran Diponegoro No. 71, Kenari, Senen, Jakarta Pusat 10430.

## Abstract

**Introduction:** Thalassemia is an inherited hemoglobinopathy with high prevalence and incidence in Indonesia. It leads to health, psychosocial, and economic burdens that affect patients, caregivers, and the country. As treatments for thalassemia in Indonesia remain expensive, not readily available, or associated with poor compliance, thalassemia prevention through screening programs is highly recommended to reduce the number of new cases. It is best to target thalassemia prevention and education to youth, but baseline data regarding their knowledge, attitude, and practice must first be assessed to measure their current awareness level as well as behavioral patterns regarding thalassemia. Currently, there has been limited research on public perception towards thalassemia in Indonesia.

**Methods and analysis:** This observational, cross-sectional study will recruit at least 500 participants between the age of 15 - 24 across all provinces in Indonesia. This is the first nationwide thalassemia study that explores knowledge, attitude, and practice among Indonesian youth (15 - 24 years old) – including thalassemia major patients, carriers, unaffected individuals, and individuals with unknown carrier status. A questionnaire will be disseminated online through social media. The questionnaire will consist of 28 questions to assess knowledge, attitude, and practice from the general population and 38 questions to assess knowledge, attitude, and practice specifically from thalassemia major patients. Questions about whether thalassemia is perceived as a curse, role of consanguinity on the mode of inheritance, and willingness to undergo screening are incorporated to specifically suit Indonesian sociocultural settings.

**Ethics and dissemination:** This study has been approved by the Ethical Committee of Faculty of Medicine, Universitas Indonesia, and Cipto Mangunkusumo Hospital. Informed consent will be obtained from all participants before completing the online questionnaire. Results will be published in a relevant journal and scientific meetings as well as shared with local stakeholders and policymakers.

**Study registration:** This study has been registered at ClinicalTrials.gov (NCT04706585).

**ARTICLE SUMMARY:** *Strengths and limitations of this study:*

- First nationwide knowledge, attitude, and practice study on Indonesian youth that provides baseline data for thalassemia major patients, carriers, unaffected individuals, and individuals with unknown carrier status.
- Incorporation of knowledge, attitude, and practice questions that are specific to Indonesian cultural settings, which include perception of thalassemia as a curse and impact of consanguinity on inheritance of thalassemia.
- Online dissemination of the survey via social media allows us to reach a large number of Indonesian youth from all backgrounds who are located in all 34 provinces of Indonesia
- The use of online self-administered questionnaire for data collection may discourage those with poor Internet connection, particularly in remote areas, from participating.
- Lack of background data on public perception towards thalassemia in Indonesia means that some assumptions from expert opinions and educated guesses have to be made when developing the questionnaire.

## INTRODUCTION

Around the globe, it is estimated that around 330,000 people are affected by hemoglobin abnormalities (hemoglobinopathy) every year, and thalassemia is a type of inherited hemoglobinopathy that is responsible for most of the cases[1]. Thalassemia is a condition where mutation occurs to the globin gene, which causes a defect in the α-globin or β-globin hemoglobin chain. This results in α-thalassemia and β-thalassemia, respectively[2]. The most severe type is β-thalassemia major as the patients will have severe anemia and require lifelong blood transfusion[1]. A significant number of thalassemia cases are found within the “thalassemia belt”, which spans across the Mediterranean ocean to Asia, including Indonesia. Approximately 5% of Indonesian population carry the thalassemia gene as carriers. With a total population of 270 million people, this percentage is really high when compared to that in other countries within the thalassemia belt[3,4]. Moreover, the data from 2019 showed that Indonesia has around 10,500 diagnosed cases across the country, and it is projected that the number could reach 20,000 by 2020. Nevertheless, these numbers are likely underestimated since many thalassemia cases in Indonesia remain undiagnosed[5,6].

In addition to high prevalence and incidence in Indonesia, thalassemia carries a high burden on the lives of patients. Thalassemia major patients are required to undergo blood transfusion as one of the therapies. However, long-term and routine blood transfusion may result in iron overload on organs such as the heart, liver, endocrine organs, and bone[7]. In addition, thalassemia major patients may suffer from conditions such as impaired growth and development, delayed puberty, mental retardation, depression, and anxiety[8-10]. This may lead to lower productivity as studies showed that thalassemia patients are frequently absent from school or work. Around 34% of patients on employment were absent from work for one or more days, while around 40% of students were absent from school for more than 8 weeks[11,12]. Among patients from the youth age group, poor psychosocial wellbeing is commonly found as stunted growth, skeletal abnormalities, and delayed puberty cause dissatisfaction towards body image[13,14]. Since thalassemia is a chronic condition, it also exposes caregivers to high stress levels. Overall, thalassemia negatively impacts the quality of life of patients and their caregivers[15].

It is possible to reduce the number of thalassemia cases through adequate prevention, screening, and education programs. Several countries within the thalassemia belt such as Cyprus, Greece, and Italy already implemented thalassemia prevention programs to reduce the number of thalassemia cases in their countries[16]. By 2007, Cyprus had successfully cut down the number of thalassemia major cases into zero cases following a mandatory national thalassemia prevention program which includes screening and prenatal diagnosis[17]. Similar preventive approach may even offer greater benefits to low-to-middle-income countries including Indonesia, where definitive treatments (i.e., bone marrow transplant, hematopoietic stem cell therapy) and supportive treatments (i.e., blood transfusion, iron chelating drugs) for thalassemia remain either expensive, not readily available, or associated with poor compliance[6,18]. The Indonesian Ministry of Health claimed that funding for thalassemia treatment accounted for USD 149 million between 2014 and 2018, putting it as the fifth most expensive spending from the total healthcare budget. Treatment for thalassemia patients in Indonesia can cost up to USD 28,500 per person, whereas the budget for thalassemia screening only cost USD 29 per person[19]. Hence, in addition to reducing the number of newborns affected by thalassemia every year, prevention and screening for thalassemia can also save a country from high economic burden. As stated by Indonesian Ministry of Health, screening for thalassemia aims to identify carriers and inform them about the chances of having a thalassemia-affected offspring as well as the preventive measures. To date, only Jakarta, the capital city of Indonesia, mandates mean corpuscular volume (MCV) and mean corpuscular hemoglobin (MCH) examinations prior to marriage[6]. It is currently unclear whether the Indonesian government plans to extend this policy about mandatory premarital screening to other regions[20].

Despite the good intention of premarital screening policy, the majority of couples screened may still proceed to marriage regardless of their screening result. This often happens in highly religious countries including Indonesia, where the legal dimension of marriage is intertwined with cultural and religious values. A study in Iran discovered that half of all couples proceeded to marriage despite the fact that they were carriers of β-thalassemia trait[21]. Another study in Central Java, Indonesia, found that half of the couples who applied for marriage had poor knowledge regarding thalassemia and none of them participated in premarital screening[20]. To overcome this limitation, some countries have suggested the implementation of antenatal screening, which is performed during pregnancy[16]. However, since Indonesian law currently does not allow abortion following antenatal screening, antenatal screening is not a feasible solution. This dilemma has led experts in Indonesia to suggest that it may be best to conduct thalassemia screening programs on school-aged children and university students, so they can consider potential marriage partners from an early age[6].

In order for prevention and education programs for the youth to be successful, the level of their knowledge, attitude, and practice (KAP) must first be assessed. This is because KAP often translates into awareness level towards a disease and enables behavioral patterns of a specific population to be defined. Results from past studies in other countries regarding KAP towards thalassemia have been used to inform thalassemia screening policy and education program[22-25]. Considering the high burden of thalassemia as well as lack of prevention and education programs in Indonesia, there is an urgent need to investigate KAP that is representative of the Indonesian youth population.

## METHODS AND ANALYSIS

### Study design and aims

The study is designed as an observational, cross-sectional survey using a newly developed questionnaire. The primary objective of this study is to assess the knowledge, attitude, and practice of young people in Indonesia towards thalassemia. Another objective is to determine their willingness to undergo thalassemia screening independently. Results from the study will also be intended to serve as a foundation for future research and policies to inform better thalassemia prevention and promotion strategy.

### Time period

The study started its planning phase between March 2021 and April 2021 and is expected to run until August 2021. Questionnaire distribution to collect the data will begin in June 2021.

**Table 1.**
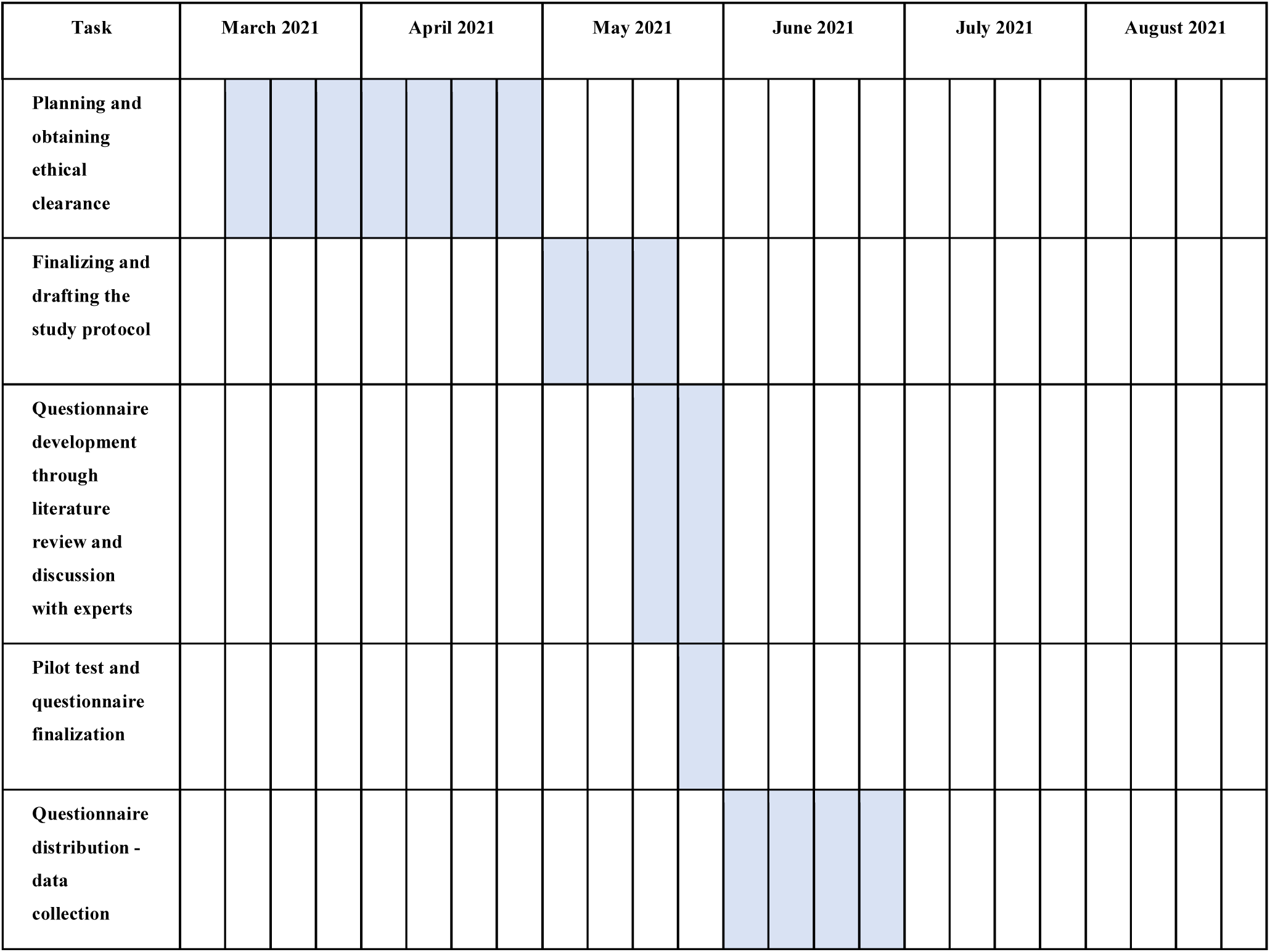
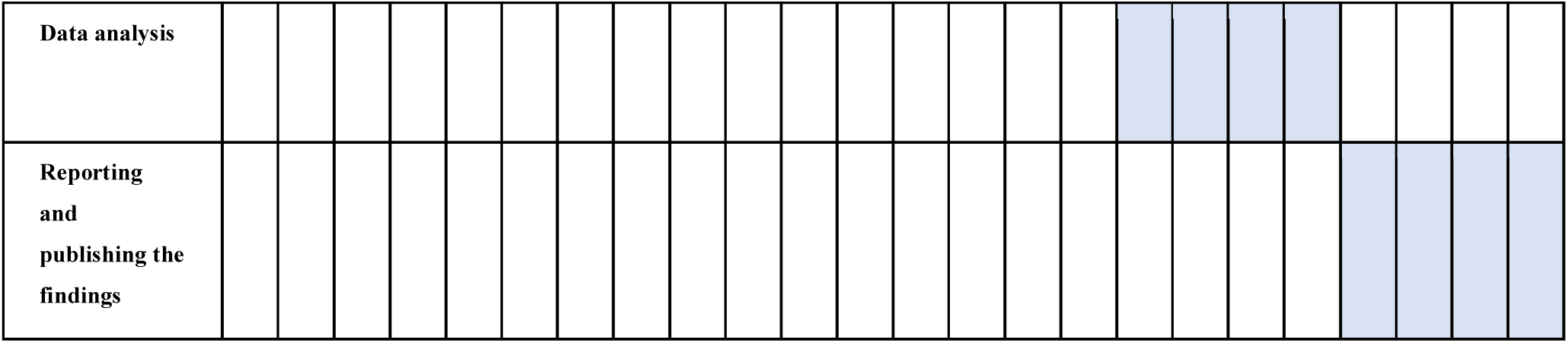
Gantt chart depicting the study timeline.

### Study settings

This study is developed as a nationwide study conducted in Indonesia. Indonesia is located in Southeast Asia and exists as the largest archipelago (17,508 islands) in the world. It has been reported that there are more than 600 ethnic groups and 6 official religions in Indonesia. Javanese is the largest ethnic group, and most Indonesians identify themselves as Muslim. According to the latest figures from The World Bank and United Nations, Indonesia is the fourth most populous country in the world with a population size of 275,064,108. The majority of its population, however, is concentrated in Java island[4,26]. The youth constitute a significant portion of the total population. In a report by the United Nations Population Fund (UNFPA), it was stated that around 26% of the Indonesian population belonged to the 15-29 age group in 2010, and by the end of 2035 an addition of 70 million youth is projected[27]. In terms of economy, Indonesia has recently been promoted to an upper middle-income country after being a lower middle-income country for more than two decades.

Due to the ongoing pandemic, the questionnaire used in this study will be online-based using Google Form. Google Form is the chosen platform as it is free and widely used in the country. Moreover, it offers a variety of question types that suit this study’s aims (e.g., Likert scale, multiple choice, checkboxes, drop-down).

### Study population/Eligibility criteria

Since the scope of the research is nationwide, this study aims to recruit a representative sample of the Indonesian youth population. Young people will be eligible to participate if they meet all of the following inclusion criteria: 1) Willing to provide informed consent after reading and understanding the information letter; 2) Within the age range of 15-24 years. Participants who are currently not residing in Indonesia will be excluded. To define who is included in the youth age group, an operational definition from the United Nations was used (“*young people between the age of 15 and 24 years*”)[28]. Thalassemia major patients, carriers, unaffected individuals, and individuals with unknown carrier status will all be enrolled.

### Sample size calculation

The minimum sample size is calculated using the formula: N = (Zα^2^ x p x q) / L^2^. Zα is the level of confidence, p is the probability of individuals that have the disease, q is the probability of individuals who do not have the disease (1 - p), and L is the precision of estimate or the margin of error. Value for Zα is 1.96 (95% confidence interval), p is 0.5, and q is 0.5. The value 0.5 is chosen for p since reliable data on the number of thalassemia patients in Indonesia, despite its high prevalence, is not available. Value for L is 0.10 for thalassemia major patients and 0.05 for the rest of the participants. The result of the calculations is 480. This is rounded up to the nearest hundred, so a minimum of 500 participants is required for this study.

### Development and refinement of questionnaire

#### 1. Stage 1 - Translating and drafting the questionnaire

Since there was no previous study in Indonesia that corresponds to our research objectives, a self-developed questionnaire is produced. Literature search is conducted through Google Scholar, PubMed, and other databases which include studies that investigate KAP for thalassemia in other countries. The researchers, who have proficient English language skills, then compile together a list of previously used and validated KAP questions and translate them into Bahasa Indonesia, which is the official language of Indonesia. Questions that are suitable with the context of Indonesian population will be given priority. The list of questions will then be discussed and appraised together with a professor of pediatric hematology involved in thalassemia management for many years and an expert physician with expertise in public health research. A combination of 12 questions for knowledge, 10 questions for attitude, and 6 questions for practice is selected from the list to assess the KAP of members from the general population. Furthermore, an addition of 8 specific questions for knowledge and 2 specific questions for attitude is also added but only to be answered by thalassemia major patients only. Hence, there will be a total of 28 questions to assess KAP of members from the general population and 38 questions to assess KAP of thalassemia major patients.

The questions will be in multiple choice format for the knowledge component (Yes/No/Don’t know and statements to choose) and four-point Likert scale (1 = fully disagree; 4 = fully agree) for attitude and practice components. KAP questions about thalassemia include its cause, mode of transmission and inheritance, carrier, signs and symptoms, therapy, complications, prevention and screening, and information-seeking behavior.

In addition, the questionnaire includes questions about the participants’ sex, age, ethnicity, religion, location, socioeconomic level, education level, parents’ characteristics, source of information for thalassemia, and other demographic information. All questions will be uploaded into one single Google Form and made online. Through a customizable feature in Google Form, the additional 8 knowledge and 2 attitude questions can only be seen and answered by thalassemia major patients but not by the rest of the participants.

#### 2. Stage 2 - Pilot test and final revision of questionnaire

The questionnaire will be piloted among 50 randomly chosen eligible participants to evaluate its difficulty, readability, validity, and reliability. These selected participants will be asked whether they believe that some questions are hard to comprehend or not appropriate. After re-evaluation of feedback by the research team and same panel of experts, changes will be made where necessary. These may include modification of the question wording, deletion of inappropriate or irrelevant questions, or addition of questions. The participants who are asked to complete the questionnaire during pilot test will not be included in the whole study sample.

**Figure 1.**
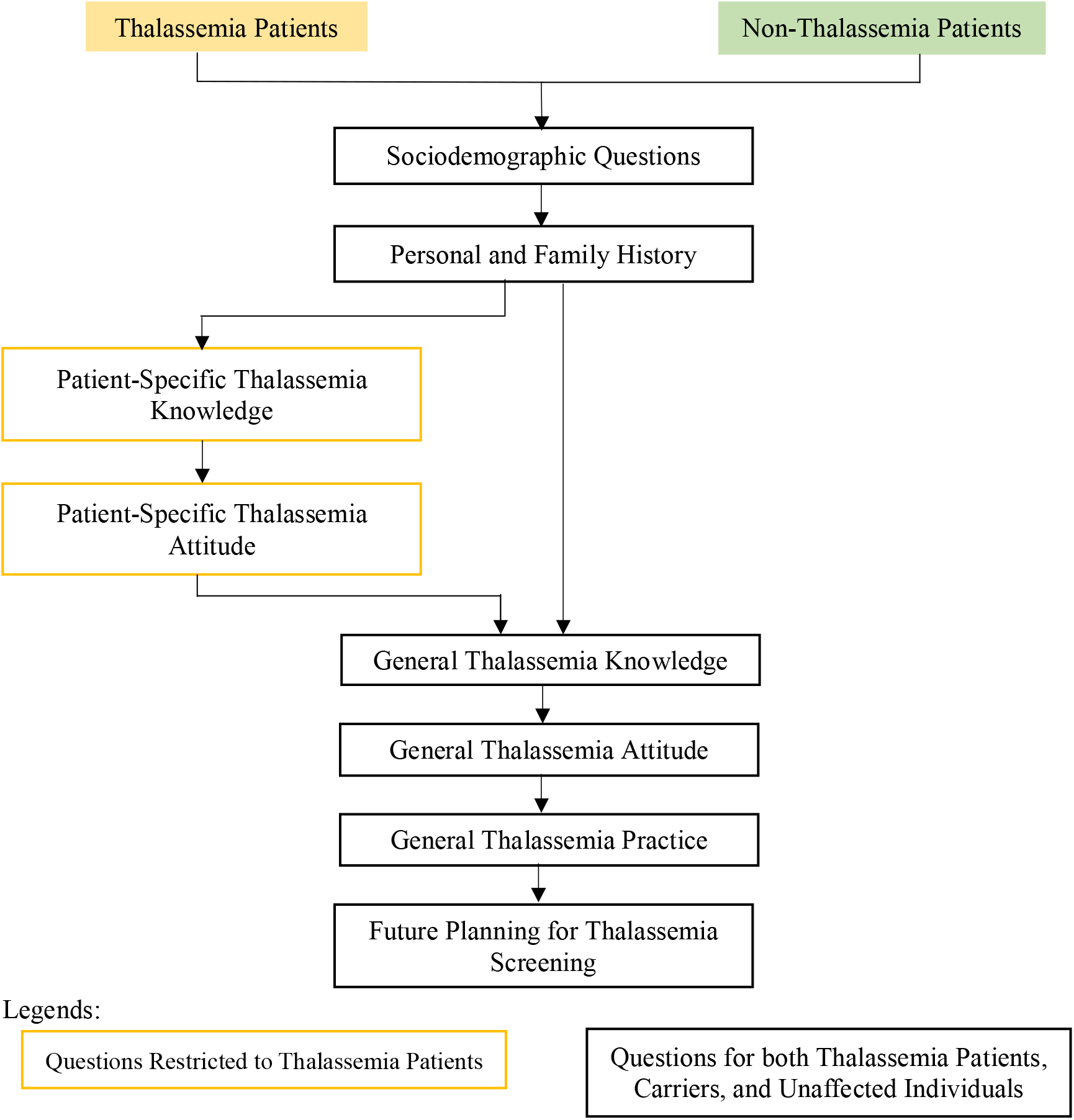
Flow chart of the questionnaire.

### Informed consent

After reading and understanding the research aims, potential participants will be asked to give their consent in the first page of the questionnaire. They cannot proceed to the next section without consent. There will be no attempt to coerce potential participants to give consent, and they are free to withdraw anytime during the questionnaire completion.

### Questionnaire distribution and coordination

This study uses convenience and snowball sampling methods. Starting from the first week of June 2021 until the end of the month where necessary, the online survey link will be distributed mainly through social media. The social media platforms include Facebook, Twitter, Instagram, WhatsApp, and LINE as they all have a large share of audience from Indonesia. Visual aid in the form of posters will also be produced and deployed to help attract attention. Each member of the research team is responsible for distributing the link and poster with as many peers and colleagues as possible. Every week, a virtual meeting will be held to evaluate the progress and number of responses. If responses from a particular province or demographic group are lacking, the research team will encourage participants from those groups to distribute the link with their network. For example, if there is still a limited number of responses from middle school and high school students, the research team will enlist the help of teachers and schools. Another targeted approach for recruiting thalassemia patients will be to work with youth groups and non-governmental organizations (NGOs) specializing in thalassemia to disseminate the survey link.

### Data management

After the data collection period, all responses in Google Form will be downloaded as Microsoft Excel file. Data will be cleaned to exclude duplicates and invalid responses. Responses from thalassemia major patients and the rest of the participants will then be put in two separate documents as they are intended to be analyzed separately. All files will be encrypted with a password that can only be accessed by the research team. Finally, the data sets will be transferred to SPSS version 24 software for statistical analysis.

### Data analysis

For KAP assessment, the widely adopted Bloom’s cutoff points are the following: 80-100% (good KAP), 60-79% (moderate KAP), and less than 60% (poor KAP)[29-31]. In this study, a modified Bloom’s cutoff value of 75% will be used to categorize participants’ knowledge, attitude, and practice into two levels: poor/negative and good/positive. This cutoff value is also based on previously published KAP studies[32-35].

Regarding the knowledge component, the number of correct answers by each participant will be automatically scored by Google Form based on our answer key. A correct answer will be given 1 point while an incorrect or “don’t know” answer will be given 0 point. For thalassemia major patients, a score from 0 to 14 indicates “poor” knowledge, whereas a score from 15 to 20 indicates “good” knowledge. For the rest of the participants, a score from 0 to 8 indicates “poor” knowledge, whereas a score from 9 to 12 indicates “good” knowledge.

Regarding the attitude component, a total score based on the responses to statements on a four-point Likert scale will first be computed. For thalassemia major patients, a score from 12 to 35 reflects “negative” attitude, whereas a score from 36 to 48 reflects “positive”. For the rest of the participants, a score from 10 to 29 reflects “negative” attitude, whereas a score from 30 to 40 reflects “positive” attitude.

Regarding the practice component, a total score based on the responses to statements on a four-point Likert scale will first be computed. Since the set of questions for practice is identical for both thalassemia major patients and the rest of the participants, the score range used is the same. A score from 6 to 17 reflects “poor” practice, whereas a score from 18 to 24 reflects “good” practice.

The result of descriptive statistics will be analyzed to assess the overall trend in KAP scores and summary of the sample demographic characteristics. Moreover, responses towards willingness to undergo thalassemia screening and information source related to thalassemia will be described. Chi-square test will be used to evaluate the association between participants’ KAP levels and their demographic characteristics, followed by logistic regression to determine the relative importance of certain variables in predicting participants’ KAP levels. Odds ratio (OR) and 95% confidence intervals (CI) will be calculated. Pearson correlation will be used to assess whether knowledge, attitude, and practice scores are associated with one another. Significance level is set at p < 0.05.

### Ethics and dissemination

The study has been granted ethical clearance by the Ethical Committee of Faculty of Medicine, Universitas Indonesia, and Cipto Mangunkusumo Hospital (KET-1517/UN2.F1/ETIK/PPM.00.02/2020). Prior to questionnaire distribution, any necessary changes in protocol will be communicated to and approved by the same ethical committee. Informed consent will be obtained from all participants before completing the online questionnaire. Results will be published in a relevant open-access, peer-reviewed journal and scientific meetings as well as shared with local stakeholders and policymakers.

### Confidentiality and data retention

The responses collected through the online questionnaire are meant to be confidential and anonymous. Participants will not be asked to include their name, address, or other personal information that may be traceable. The only exception is phone numbers, in which participants have to include it in the early section of the questionnaire. This, however, is not part of the study’s interest and will only be used as participant’s unique identifier to remove duplicate responses. It will not be used for other purposes or published anywhere. There will be no attempt to identify the individuals to which data belong to. All collected data will be stored in a cloud storage that can only be accessed by the research team. Data backup will also be made and stored in the primary investigator’s personal computer.

### Patient and public involvement

Both thalassemia patients and the general public are involved in the conception of the study, development of the study protocol, finalization of the questionnaire, and sample collection procedure. They will also be involved in dissemination of the key findings from our study, as short campaign and webinar will be planned to raise thalassemia awareness among Indonesian youth. Feedbacks and personal experiences from thalassemia patients and their community are gathered to inform the study’s background and research questions. Prior to widespread survey distribution, the questionnaire will be piloted among 50 young people in order to suggest changes in the questionnaire content. Lastly, all participants can provide criticism and suggestions as the phone number of a representative of the research team will be attached in the front page of the online questionnaire.

## DISCUSSION

Despite the notion that prevention is fundamental in reducing the high disease burden of thalassemia, most existing studies focused on the clinical aspects of thalassemia[36-39]. On the other hand, public perception and awareness towards the disease, especially in Indonesia, remain understudied. To our knowledge, this is the first nationwide study that explores KAP towards thalassemia among Indonesian youth. Currently, 80% of the productive age population in Indonesia (15-64 years old) are mainly from the reproductive age group (15-49 years old)[40]. This study focuses on Indonesian youth of age 15-24 as they will soon enter the stage of marriage and parenthood, since thalassemia gene can be passed down from parents to their offspring. Previously, a study in Semarang, Indonesia explored the KAP of 96 medical students towards thalassemia prevention. Based on the study, a large proportion of the respondents had good knowledge (42.7%), positive attitude towards thalassemia screening (72.9%), good practice towards preventive measures of the disease (91.7%), but lacked the willingness to undergo screening immediately[41]. However, its results are not representative of the whole Indonesian youth population as the study was targeted to medical students in one region.

This study includes thalassemia major patients, carriers, unaffected individuals, and individuals with unknown carrier status as it wants to test the hypothesis about whether thalassemia major patients have better KAP than the rest of the respondents. It is assumed that thalassemia major patients are equipped with better knowledge about the disease as they have undergone series of counselling with their healthcare providers. Education about the disease is an essential component of a patient’s life as it can promote their capability to perform self-care[42]. In correlation to this, the study also aims to measure to what extent thalassemia major patients understand their disease.

Considering the ongoing pandemic in Indonesia, the questionnaire will be disseminated online. A major advantage of using an online questionnaire is that all responses are automatically gathered in one online sheet. The researchers will be able to observe real-time demographic distribution of the respondents. Hence, if a particular region or demographic group is underrepresented, targeted questionnaire distribution through snowball sampling can be performed immediately. Online distribution of the questionnaire also allows for a streamlined data collection approach, faster data collection, easier monitoring, and less resources. Throughout the pandemic, this method has gained increasing popularity among observational, cross-sectional studies.

Regarding knowledge and attitude in Indonesian context, the study specifically incorporates questions about whether thalassemia is perceived by respondents as a curse. This is to explore whether such misguided stigma, as what is perceived by the public towards leprosy, is also present towards thalassemia. A study that explored public perception towards leprosy in Cirebon, Indonesia revealed that some patients believed that sorcery is one of the causes of the disease. In practice, these patients also prefer seeking treatment from traditional healers instead of licensed physicians[43].

As thalassemia is inherited through autosomal recessive manner, consanguineous marriage increases the risk of passing down the gene to offspring[44]. The prevalence of incest cases in Indonesia has increased over the decade. Surprisingly, 77% of the perpetrators are biological fathers or step-fathers of the daughter[45]. Hence, the questionnaire also includes questions that explore respondents’ view towards consanguinity and its relation to thalassemia.

Furthermore, the questionnaire also explores respondents’ willingness to undergo thalassemia screening and when do they plan to do so. A question that measures the proportion of respondents who have had screening before is also included. The decision to include these questions is influenced by previous studies that suggested there is a high prevalence of thalassemia carriers across several areas in Indonesia[46-50]. It is important to understand the trend of the answers to these questions in order to inform future thalassemia prevention, screening, and education programs in Indonesia.

Despite the study’s strengths and novelty, there are several challenges that may be encountered during the data collection process. Since the questionnaire will be fully online, respondents in remote locations with poor Internet connection may not be able to access it. Additionally, as the questionnaire will be self-administered, respondents who are struggling with functional illiteracy may also have difficulty in understanding the questions. Last but not least, the findings of our study may not be generalizable to populations of other age groups and other countries.

## CONCLUSION

High burden of thalassemia in Indonesia can be significantly reduced through adequate prevention, screening, and education programs. These programs are best delivered to the youth population who will soon enter the stage of marriage and parenthood, so they can consider potential marriage partners from an early age and therefore prevent the birth of newborns with thalassemia major. Through the use of an online questionnaire, this novel study will measure the KAP towards thalassemia among Indonesian youth. Several questions that are unique to the Indonesian settings such as whether thalassemia is perceived as a curse and the role of consanguinity in its mode of inheritance are included. The results of this nationwide study will serve as the foundation for future studies and policies.

## Data Availability

All data for this manuscript is available upon request to the authors

## LIST OF ABBREVIATIONS

CI: Confidence interval
KAP: Knowledge, attitude, practice
MCH: Mean corpuscular hemoglobin
MCV: Mean corpuscular volume
NGO: Non-governmental organization
OR: Odds ratio
SPSS: Statistical Package for the Social Sciences
UNFPA: United Nations Population Fund
USD: United States dollar

## Acknowledgement

We are grateful to Thalassemia Movement community for their help in disseminating the online survey particularly to thalassemia major patients across Indonesia. We would also like to express our gratitude to the Faculty of Medicine, Universitas Indonesia for the support.

## Authors contribution

MMW, ECY, VRT, MY, and PAW equally contributed to the conception of the study, development of the study protocol, and development of the questionnaire. MMW, ECY, and VRT performed literature search and drafted the manuscript. MY and PAW reviewed the manuscript and suggested necessary changes. All authors have approved of the final version of this manuscript.

## Funding statement

This research received no specific grant from any funding agency in the public, commercial or not-for-profit sectors.

## Competing interests statement

The authors declare that they have no competing interests..

## Notes

### Competing Interest Statement

The authors have declared no competing interest.

### Clinical Trial

NCT04706585

### Funding Statement

No external funding recieved

### Author Declarations

Ethical Committee of Faculty of Medicine, Universitas Indonesia, and Cipto Mangunkusumo Hospital (KET-1517/UN2.F1/ETIK/PPM.00.02/2020).

### Summary of Updates

Simplified ClinicalTrial.gov registry information; Change author list; Change Gantt chart to address postponement of recruitment to June-July 2021; Change anticipated recruitment date to June 2021

